# An Exploratory study for characterIzing and predicting prostate abNormalities uSing MRI-based radiomics and arTificial intEllIgeNce (EINSTEIN)

**DOI:** 10.1101/2023.11.26.23299017

**Authors:** Yassine Bouchareb, Gayathri Delanerolle, Yarab Al-Bulushi, Ali Al-Khudhuri, Srinivasa Rao Sirsangandla, Ghalib Al Badaai, Heitor Cavalini, Peter Phiri, Ashish Shetty, Jian Qing Shi

## Abstract

**Introduction:** Prostate cancer (PCa) is the fourth most prevalent cancer globally, and the most common among men. Most PCa patients in Oman are presented during the advanced stages of the disease with widespread metastatic disease reducing their overall rates of survival. Characterisation of the Omanis PCa population could be beneficial to develop a clinical profile demonstrating specific characteristics to better classify and derive radiomics signatures. These could help in developing artificial intelligence methods to assist with earlier and quicker diagnosis of possible prostate lesions.

**Methods:** A retrospective, cross-sectional study has been designed to determine the pathological and radiological characteristics based on multi-sequence 3-dimensional Magnetic Resonance Imaging (MRI). The MRI records are maintained within the existing electronic healthcare records of the Sultan Qaboos University Hospital’s Department of Radiology and Molecular Imaging. Data will be extracted based on a confirmed diagnosis reported between the 1^st^ of January 2010 and October 2023. All patients included within the study will be aged between 18-99 years. A study specific data extraction template has been devised to gather demographic details, clinical parameters and radiological findings based on existing imaging reports within the HIS and PACS systems.

**Ethics approval:** Research Ethics approval reference for this study is (MREC #3176 REF. NO. SQU-EC/ 283\2023)

**Conclusion:** The data analysis will be conducted using statistical software to conduct a Joinpoint regression analysis and linear regression modelling. We will also conduct a descriptive analysis.

## Background

Prostate cancer (PCa) is the most common cancer next to the skin cancer in men. It poses a substantial burden on the public all over the world. The incidence of overall (PCa) is increased by 3% per year since 2014, and it is 5% for advanced (PCa) [1]. It is a most leading cause of death in men and it can be heritable partially. Depending on the pathological risk, PC associated mortality varies with a factor of 10 in the category [2]. (PCa) mortality can easily be reduced with early detection and primary diagnosis. In Arab countries, the most of the PC patients showed advanced stages with distance metastases, when compared to their counterparts in the USA [3]. In Oman, PC is the most common cancer affecting Omani men. Its crude incidence is 6.6 and age-standardized rates (ASIR) of 12.2 per 100,000 of population per year [4]. The incidence is expected to rise due to increased life expectancy in Oman and better health standards. A recent report showed that more than 70% of PC patients present with advanced stages of cancer (C & D) [5]. The ultrasound-guided transrectal (TRUS) prostate biopsy with up to 12 cores used to be the standard method of diagnosing PC for many decades. Later, the Gleason score was developed, in which the tumor was categorized in low-risk, intermediate-risk, and high-risk (PCa) [6]. Due to various disadvantages, transrectal ultrasound examination along with contrast enhanced sonography, ultrasound elastography, or computer guided sonography were not recommended for the primary diagnosis of PC [7]. Currently, MRI with high-resolution images with three different contrasts, or MRI Sequences (hence “multiparametric“) is routinely in clinical practice for the screening of PC [8]. In recent years, distant metastases detection and identifying the risk of recurrences have been significantly improved with advanced developments in imaging techniques [9]. The prostate-specific antigen (PSA) plays a significant role in (PCa) diagnosis. It was estimated that PSA screening contributed to an almost 50% reduction in (PCa) mortality [10]. Nevertheless, PSA usage is known to be associated with overdiagnosis as well as overtreatment of non-aggressive (PCa) [11]. Although new serum- and urine based diagnostic tests, genetic tests, and advanced imaging modalities have great potential to reduce the risk of overdiagnosis, obtaining the abundant data from these new tests is a major clinical and research challenge [12].

The multiparametric MRI (mpMRI) and MR/TRUS fusion biopsy (TRUS = transrectal ultrasonography) investigations reduced the overdiagnosis of low-risk PCa and increased the specificity of initial detection of (PCa) and assessment of the prognosis. Prostate-specific membrane antigen (PSMA) hybrid imaging is helpful in identifying the lymphogenic and bony metastases in patients with high-risk (PCa) [8]. Rare pathogenic mutations (RPMs) were known to be crucial in recognizing the PC risk. Mutations DNA damage repair (DDR) genes including homologous recombination such as BRCA1, BRCA2, and ATM or mismatch repair genes such as MLH1, MSH2, and MSH6) are associated with PC risk. Additionally, variations of HOXB13 gene which is linked to urogenital development are strongly associated with PC risk [13].

Artificial intelligence (AI)-based systems could be another potential option for addressing these challenges. In addition to the clinically relevant parameters, advanced radiomics using standardized quantitative feature extraction methods (Image Biomarker Standardization Initiative [14]), machine learning and deep learning methods will be employed to extract and characterize specific patterns of prostate lesions based on T1, T2 and Diffusion Weighted MR imaging (DWI) [15].

## Methods

We designed a retrospective study to determine the primary imaging features and clinicopathological characteristics of prostate lesions in Omani patients. This study will be looking specifically at Prostate imaging that is performed in multi-sequence magnetic resonance imaging (MR) of patients who visited the Department of radiology and molecular imaging, Sultan Qaboos University Hospital during the period January 2010 to October 2023.

### Inclusion criteria

Male adults that underwent Prostate MRI in Sultan Qaboos University Hospital from January 2010 to October 2023 and for whom a biopsy was performed.

MRI technique for prostate cancer consist of routine protocol multiplanar and multi-sequential MR images of the pelvis focused on the evaluation of the prostate in 1.5-3 Tesla scanner machines. High resolution T1 and T2 images of the prostate are obtained in axial, coronal and sagittal planes. Diffusion weighted images of the prostate performed in the axial plane. Post-contrast dynamic evaluation of the prostate with delayed FAT SAT T1 weighted images after administration of Gadolinium in the axial plane. Imaging is conducted with the patient in supine position using a surface coil. Fasting for at least 4 hours was recommended to reduce bowel artefacts.

*Exclusion criteria:* Suboptimal prostate MRI scans due to variable reasons such as excessive motion, inadequate field of view or missing critical sequences. MR of patients who underwent radical prostatectomy. Patients with no prior Prostate MRI at Sultan Qaboos University Hospital.

### Primary objectives

1. Describe and identify the most common clinical and imaging characteristics of prostate cancer patients in a large Tertiary Care Hospital in Oman.
2. Characterize prostate disorders and abnormalities using radiomics on MR imaging sequences

### Secondary objective

1. Classification and deriving radiomics signature of prostate cancer lesions using Machine learning and Deep learning methods.

### Data collection

A sample size of approximately 300 patients will be secured based on biopsy and MRI data available within the electronic healthcare records dated from the 1^st^ of January 2010 to 1 ^st^ o f October 2023 and their clinical and pathological parameters will be included in the study. The data collection will be completed using a study specific extraction template that will include demographic information such as age, race, ethnicity, region, height, weight, BMI, family history, risk factors like hypertension, diabetes, smoking history, hyperlipidemia, chronic kidney disease, ischemic heart disease, and other malignancies. Clinical parameters (Onset of the symptoms, first visit to urology clinic, PSA level prior to MRI, history of biopsy and type of biopsy, histopathology findings with Gleason score, surgical interventions, histopathology staging, complications either both prior to intervention and after intervention, findings on other imaging evaluation such as bone scan, medications, chemotherapy, and radiation therapy). Prostate Imaging findings (Prostate size in three dimensions, findings of benign prostate hypertrophy, prostate lesions and their numbers, size and site, P-IRADS scoring for each lesion, local staging and invading of adjacent structures like seminal vesicles and neurovascular bundle, adjacent suspicious lymph nodes and distal metastasis).

Additionally, manual contouring of each prostate observation will be performed using a dedicated software package. The volumetric data will be utilized for radiomics analysis for two-dimensional and three-dimensional feature extraction.

### Data analysis Radiology

Standardized tools such as 3D-Slicer and PyRadiomics [14] will be used to contour the prostate anatomical regions, prepare, extract and select radiomics features from retrospective three-dimensional MR images acquired using different sequences, including at least T1, T2 and DWI sequence of the pelvis. Clinical and standardized scores for prostate malignancies PRADs and Gleason scores will be combined with imaging features to achieve the above study objectives by means of training, validating, and testing the machine learning models. The study will explore the performance of several machine leaning models, including Naïve Bayes, Support Vector Machine, Bagging, Random Forest, K-nearest neighbors, Decision Tree and Ensemble Meta voting. The optimal model will be selected based on assessment metrics such as precision, recall and area-under-curve (AUC) by randomizing the training/validation, followed by testing using the testing sets. It is the intention of the research team members to consider the combination and an expansion of these models should the initial data analysis reveal lower than expected performance on clinico-pathological and extracted imaging features.

### Statistical

The data will be analyzed using the IBM Statistical Package for Social Sciences Software (SPSS) and appropriate statistical tests will be used to determine characteristics of prostate abnormalities. A Joinpoint regression analysis will be performed to report the trends in data over time, with a particular focus on identifying points at which a significant change, or “joinpoint,” occurs in the trend.

A linear regression and generalized regression modeling approaches will be used to develop the relationship between a dependent variable and one or more independent variables by fitting a linear equation to the observed data. The variables identified will be used to predict and explain the variation in the dependent variable. R-squared and are under the curve (AUC) will be used to measure how well the independent variables explain the variability in the dependent variable.

Machine learning algorithms will be used to determine the most common features of prostate lesions that should be accounted for prior to any treatment option, including surgical interventions.

## Dissemination

The findings of the study will be presented at different scientific events such as, international and national conferences, symposiums and seminars. High impact journals will be targeted for the publication of the study’s findings.

## Data Availability

All data produced in the present study are available upon reasonable request to the authors

